# Improving Adherence to electronic Patient-Reported Outcomes: *Study Protocol for a Systematic Literature Review and subsequent qualitative Interviews*

**DOI:** 10.1101/2024.06.13.24308883

**Authors:** Rebecca Mukowski-Kickhöfel, Lisa Otto, Alizé Rogge

**Affiliations:** Center for Patient-Centered Outcomes Research, Charité – Universitätsmedizin Berlin, Corporate Member of Freie Universität Berlin and Humboldt-Universität zu Berlin, Berlin, Germany; Department of Psychosomatic Medicine, Charité – Universitätsmedizin Berlin, Corporate Member of Freie Universität Berlin and Humboldt-Universität zu Berlin, Berlin, Germany

## Abstract

**Background:** Electronic patient-reported outcomes (ePROs) are frequently used to include the patients’ perspective in clinical trials and routine clinical care. In contrast to data collected in clinical trials, PRO response rates tend to decrease drastically overtime in routine clinical care, leading to missing data and questionable validity of results reported. This study aims to investigate factors influencing patient adherence to digitally collected PROs.

**Method:** The study comprises of three steps: (1) a systematic literature review (SLR) focusing on factors increasing PRO response rates, (2) focus group interviews with patients to develop criteria on how to increase response rates for real-world evidence (RWE), and subsequently, (3) an anonymous online survey to evaluate developed criteria. The SLR will follow the PRISMA guidelines. The inclusion criteria for the review encompass studies focusing exclusively on patients or people recruited via medical personnel. The literature review will include studies from various settings, encompassing publications in German or English language published within the past 10 years (2014-2024). Participants for the focus group interviews (n = 6-8/group) will be recruited via patient advisory boards or groups/organisations. The findings of the SLR and the focus group form the basis for the anonymous online survey to include more patients.

**Outlook:** The results of the study can be incorporated into the development and implementation of digital collection methods for PROs, with the aim of improving adherence to PROs and thus robustness of RWE data.

## 1. Background and Significance

Patient Reported Outcomes (PROs) and their respective measures (PROMs) are gaining increasing significance, driven in part by a paradigm shift in medicine towards value-based, patient-centered care (Gerst, 2015). By including the patients’ health-related quality of life, functioning and symptoms, early interventions can be incorporated into treatment plans leading to possibly better outcomes. So far, PROs have demonstrated value in clinical trials; for instance, suggesting that their integration might lead to increased survival rates in cancer patients (Basch et al., 2017; Gotay et al., 2008; Quinten et al., 2009). However, as clinical trials must adhere to strict study protocols often integrated into randomized controlled trials, their results lack external validity and might not be applicable to heterogenous patient groups in routine care (Foster et al., 2018).

The assessment of PROs in routine care has long been supported by healthcare professionals, patients and other stakeholders but its broad uptake and rigorous integration has yet to be achieved (Kelkar et al., 2016). Real-world evidence (RWE) aims to overcome the limitations from clinical trials by gathering information from routine care settings. In comparison to data collected in clinical trials, real-world data (RWD) have the potential to form a realistic picture of clinical care including heterogeneous patient groups as well as different motives/therapy targets in the collection of data. As of now, RWD is often criticized for a lack of robust reliability as patients might not fill out PROMs regularly. These missing data lead to results difficult to interpret, and therefore, to draw conclusions from.

A systematic literature review from 2020 showed that there is wide variation and a downward trend in patients’ response rates to PROMs over time across cohorts, registry-based studies and registries (Wang et al., 2020). So far, barriers and challenges to the implementation of PROs in real world settings have been discussed from a regulatory or physicians’ point of view (Foster et al., 2018); however, the patient perspective is often left out. Aside from the structural issues related to PROM implementation into routine care, a reason for declining response rates may lie within the PROMs themselves. Approaches to increase PRO adherence have often focused on gamification approaches (Almeida et al., 2023), but motivating patients to answer PROMs in order to gain points, achievemilestones or other forms of incentivization, might lead to a systematic bias.

To increase response rates to PROMs for RWE, patients’ preference and acceptance of measures might play a key role (Oehrlein et al., 2019). Early research suggests that the type of measure, the content appropriateness of the measure, frequencies and the severity of the illness might influence overall PROM adherence in routine care, and hence, build up to a decrease in response rates over time (Unni et al., 2023). The lack of robust, reliable data collected in RWE due to non-adherence to PROMs could rise to immense challenges in the future putting the value of PROMs, implementation efforts, and precision of statistical models at risk. The aim of this study is therefore to explore the patients’ perspective on the adherence to PROMs and to develop a list of criteria on how increase response rates for PROMs in clinical care under RWE conditions.

## 2. Study Aims

The aim of the study is to investigate factors that increase patient adherence to digitally collected PROMs over time. The project can be separated into 3 steps:

### Step 1

In the systematic literature review (SLR), we aim to identify factors that lead to an increase or decrease of patient response rates to PROMs. As many papers have beforehand discussed challenges when implementing PROMS, this SLR should solely focus on the structure and content of PROMs as influential factors for acceptance, adherence and response rates.

### Step 2

We will conduct at least one focus group with patients and/or patient representatives aiming to a) discuss the findings of the SLR and b) further define criteria that might lead to an increase of patient adherence to PROMs over time.

### Step 3

Criteria developed in step 2, will then be disseminated in a larger patient group (n = 50-100) within an anonymous online survey. Participants will receive the possibility to rate their level of acceptance/relevance to criteria developed. In addition, it will be asked whether the relevance of these criteria would also lead to increased adherence to the PROMs.

## 3. Method

### 3.1 Study Design and procedure

Initially, a systematic literature review will be conducted according to the PRISMA guidelines. Subsequently, an interview guide for a focus group will be developed based on these findings. At least one focus group interview is intended. Patient representatives are recruited for the focus group interview through email and newsletter outreach from patient organizations. Participants will be briefed on the scope of the interview and the handling of collected data. Alongside study details, participants will receive a consent form to sign beforehand. The interview will be conducted by a project employee online. The target number of participants is 6-8 patient representatives per group, with the interview set to last max. 100 minutes. Participants have the right to halt the interview at any time without providing reasons. Additionally, an anonymous online questionnaire will be conducted. This serves the purpose of including further perspectives from a larger patient population. Participants will also be recruited by sending an email to patient organizations (see figure 1 for the study flow).

**Figure 1:**
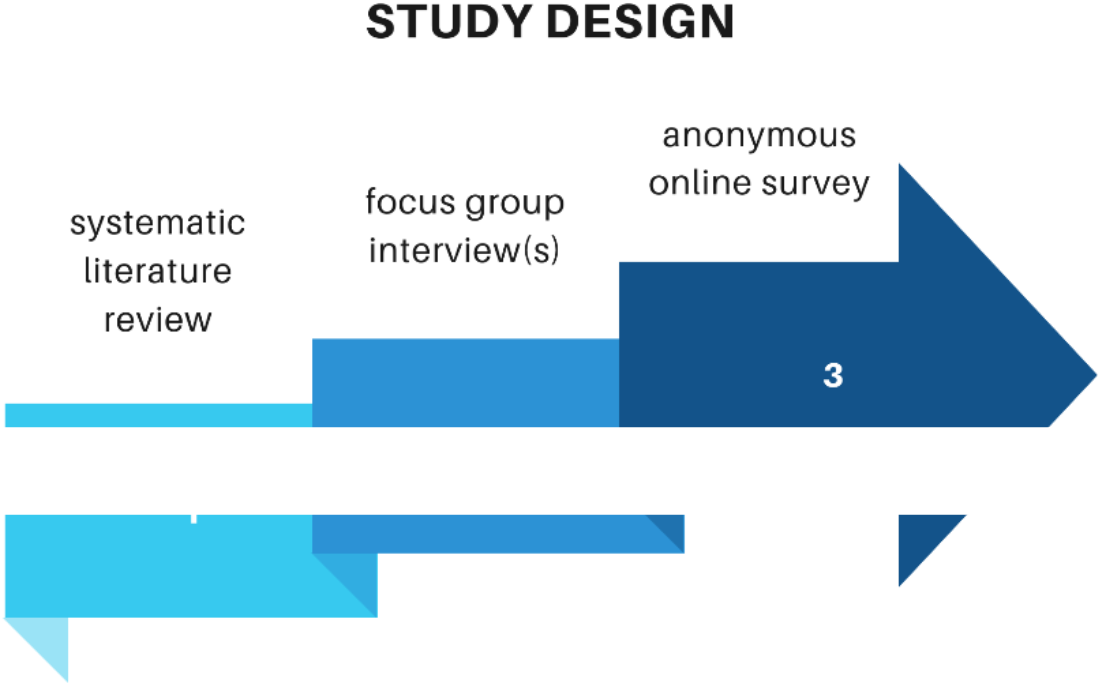
Studyflow

### 3.2 Systematic literature review

The research team will conduct a systematic literature review addressing the research question. Relevant studies will be identified through the following databases: MEDLINE, Embase [Ovid], CINAHL, PsychArticles, PsycInfo [Ebsco], Scopus and Web of Science. The review will encompass various study designs, including systematic reviews, observational studies, and qualitative studies. The inclusion criteria span studies conducted in both German and English across all settings within the past decade. The collaboration tool Rayyan (Ouzzani et al., 2016) will be utilized for the search, screening of titles and abstracts, as well as for the full-text review. The search terms illustrated in Figure 2 will be used for this purpose.

**Figure 2.**
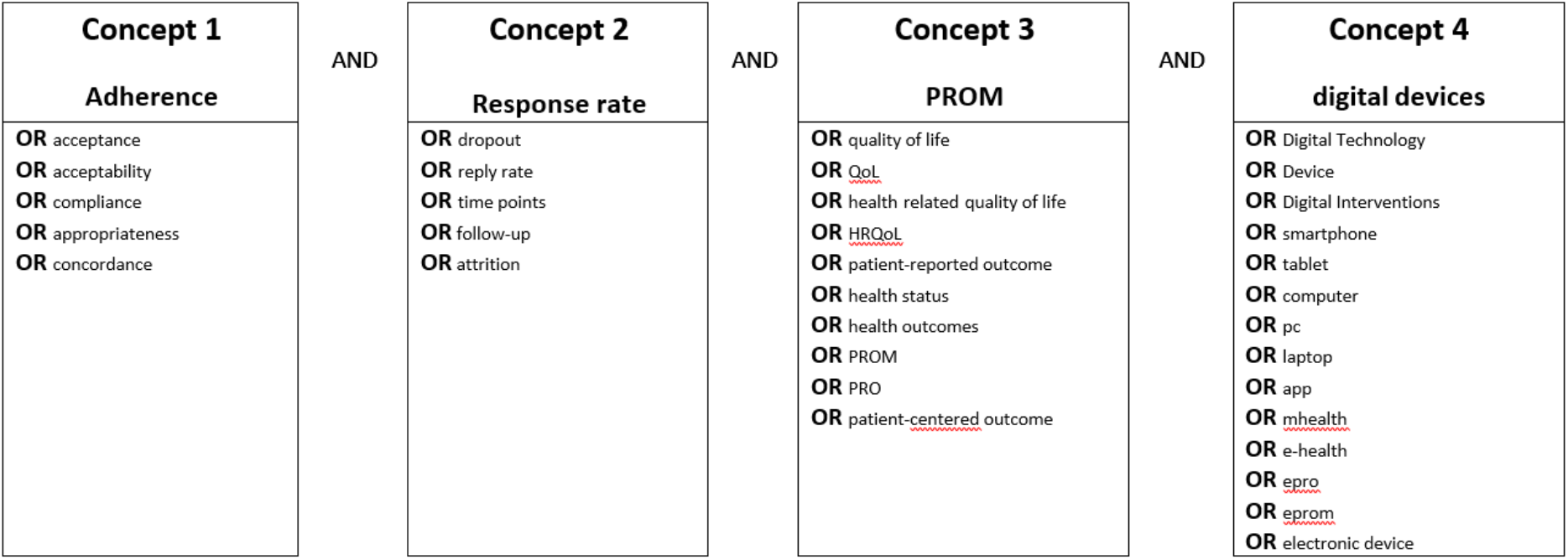
planned search terms

Following this, the research team will assess gray literature and research papers that may not have been located through the previously mentioned process. Furthermore, the reference lists of the identified papers and existing reviews will be examined to identify relevant articles, subject to the same selection process as the original papers.

Considering the inclusion and exclusion criteria outlined below, all titles will be initially screened by AR. Subsequently, the screening of abstracts will be conducted by researchers AR, LO and RMK. In the third step, the author team will make an independent selection of studies based on the inclusion and exclusion criteria. In preparation for step 2, criteria for the increase of adherence to PROs will be formulated using deductive and inductive methods following procedure introduced by Mayring et al. (2016) following the SLR results.

#### Inclusion criteria

- age ≥ 18 years
- patient
- Multiple medical contacts in routine care setting for the same disease/illness (incl. post-operative care and chronic diseases)
- Routine care setting
- Participants must be recruited via medical personnel

#### Exclusion criteria

- clinical trials (in terms of drug or intervention research)
- study protocols

### 3.3 Development of a guideline for the focus group interview

The semi-structured interview guideline used during the focus group interviews will be informed by the findings of the SLR. The following questions could be part of the semi-structured interview guideline (non-exhaustive list):

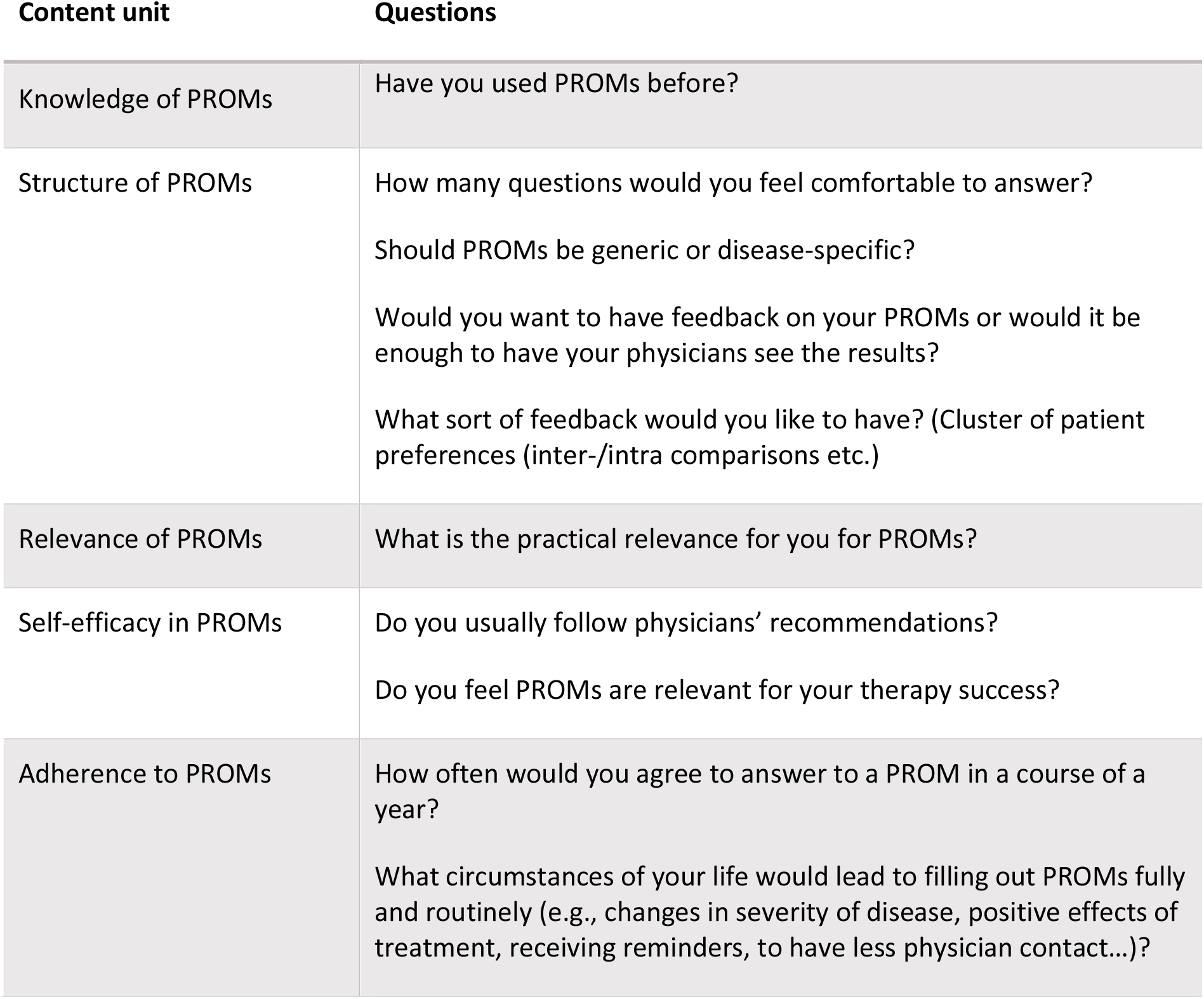

| Content unit | Questions |
| --- | --- |
| Knowledge of PROMs | Have you used PROMs before? |
| Structure of PROMs | How many questions would you feel comfortable to answer?<br>Should PROMs be generic or disease-specific?<br>Would you want to have feedback on your PROMs or would it be enough to have your physicians see the results?<br>What sort of feedback would you like to have? (Cluster of patient preferences (inter-/intra comparisons etc.) |
| Relevance of PROMs | What is the practical relevance for you for PROMs? |
| Self-efficacy in PROMs | Do you usually follow physicians' recommendations?<br>Do you feel PROMs are relevant for your therapy success? |
| Adherence to PROMs | How often would you agree to answer to a PROM in a course of a year?<br>What circumstances of your life would lead to filling out PROMs fully and routinely (e.g., changes in severity of disease, positive effects of treatment, receiving reminders, to have less physician contact...)? |

### 3.4 Focus group interview

To recruit participants for the focus group interview, several patient organizations will be contacted (see inclusion criteria above).

Interested patient representatives will receive a follow-up email containing details for the focus group, comprehensive study information, a consent form as well as an invitation link to the online interview. Before the interview begins, participants will receive oral study information. The focus group interviews will be recorded, and transcribed anonymously.

### 3.5 Anonymous online survey

Developed criteria will then be disseminated within a larger patient sample using an anonymous online survey (lime survey). Participants will be asked to rate developed criteria according to their relevance, acceptance as well as potential to increase response rates in PROMs.

### 3.6 Termination criteria

Participants in the focus group interviews and the anonymous online survey can end the study at any time without giving reasons.

### 3.7 Expected benefit

This study aims to improve RWE data collection by increasing response rates to PROMs. Moreover, results will shed further light on reasons of missing data in RWD.

### 3.8 Possible risks or burdens

There are no anticipated risks or disadvantages for the participants in the focus group interview and the anonymous survey.

## 4. Reporting the results

The results of the study will be published in a peer-reviewed journal.

## 5. Legal and ethical considerations

### 5.1 Declaration of Helsinki

The investigation will be conducted in accordance with the current version of the Declaration of Helsinki.

### 5.2 Ethics Committee

The study as described above is part of a larger research project on the implementation of PROs for research and clinical care for which an ethics approval has already been obtained (EA4/034/22).

### 5.3 Information on voluntary participation

Participation by patients is voluntary.

### 5.4 Information and consent

The study participants will be informed verbally and in written form about the nature and scope of the planned study, in particular about the possible benefits for their health and possible risks, before the start of the study. In the event of withdrawal from the study, any data already obtained will be destroyed or the patient will be asked whether they agree to the analysis of the data.

### 5.5 Data protection

The names of patients and all other confidential information are subject to medical confidentiality and the provisions of the General Data Protection Regulation (GDPR) and the State and Federal Data Protection Act (LDSG and BDSG). Personal data will not be passed on. Third parties do not have access to original documents.

Only anonymized data is collected. It is therefore not possible to delete the data of individual participants following the focus group interview or the online survey.

### 5.6 Information on insurance

No insurance is needed.

## Data Availability

All data produced in the present study are available upon reasonable request to the authors

